# EXCESS MORTALITY FROM COVID-19. WEEKLY EXCESS DEATH RATES BY AGE AND SEX FOR SWEDEN

**DOI:** 10.1101/2020.05.10.20096909

**Authors:** Karin Modig, Marcus Ebeling

## Abstract

**Objectives:** Mortality from Covid-19 is monitored in detail both within as well as between countries with different strategies against the virus. However, death counts and relative risks based on crude numbers can be misleading. Instead, age specific death rates should be used for comparability. Given the difficulty of ascertainment of Covid-19 specific deaths, excess all-cause mortality is currently more appropriate for comparisons. By estimating age- and sex-specific death rates we aim to get more accurate estimates of the excess mortality attributed to Covid-19, as well as the difference between men and women in Sweden.

**Design:** We make use of Swedish register data about total weekly deaths, total population at risk, and estimate age- and sex-specific weekly death rates for 2020 and the 5 previous years. The data is provided by Statistics Sweden.

**Results:** From the first week of April and onwards, the death rates at all ages above 60 are higher than those in previous years in Sweden. Persons above age 80 are dis-proportionally more affected, and men suffer higher levels of excess mortality than women at all ages with 75% higher death rates for males and 50% higher for females. Current excess mortality corresponds to a decline in remaining life expectancy of 3 years for men and 2 years for women.

**Conclusion:** The Covid-19 pandemic has so far had a clear and consistent effect on total mortality in Sweden, with male death rates being comparably more affected. What consequences the pandemic will eventually have on mortality and life expectancy will depend on the progression of the pandemic, the extent that some of the deaths would have occurred in the absence of the pandemic, only somewhat later, the consequences for other health conditions, as well as the health care sector at large.

## INTRODUCTION

Mortality from Covid-19 is monitored in detail both within and between countries. Even though the phrasing is often “*death rate from Covid-19*” [1], we haven’t seen any paper presenting proper death rates in the way they are usually estimated in Epidemiology or Demography, i.e. with the corresponding population at risk as the denominator with proper justification for age and sex. Doing this, would enable a more accurate picture of mortality in different age and sex strata. It would also enable comparison between men and women, and between countries with different strategies against the virus.

Neither Covid-19 death counts, nor proportion of deaths among confirmed cases are appropriate to compare between population groups within countries or between countries. This is because the underlying population structure is different, and countries differ in the way they count and define Covid-19 deaths, as well as in their testing strategies [2]. In a recent communication in the Lancet, it has therefore been argued that weekly excess deaths could provide the most objective and comparable way of assessing the scale of the pandemic and formulating lessons to be learned [3]. Deaths by all causes combined would also allow side stepping issues of what is or is not a death attributable to COVID-19 [3]. Any excess mortality in 2020 can be compared to previous years and assumed to be attributed to the Covid-19 pandemic, directly from Covid-19 deaths or indirectly through effects on other health conditions. The estimation of all-cause age-specific death rates is therefore a first step towards are more comprehensive understanding of the impact of the pandemic on the mortality of the total population.

All cause age-specific death rates provide also valuable insights if and how the pandemic dis-proportionally affects specific ages and population groups. In this context, the difference in mortality between men and women are frequently discussed. Already early during the Covid-19 outbreak in China it was observed that more men than women died from the virus. This sex difference was seen both when estimated as a proportion of infected, and when counting the number of deaths. The same pattern has since then been observed in all countries affected by Covid-19 [4], However, the comparison of mortality between men and women carry the same problems as the comparison between countries, especially since all countries have more older women than men.

For these reasons, we make use of Swedish register data about total weekly deaths, total population at risk, and estimate age- and sex-specific weekly death rates for 2020 and the 5 previous years. By this we aim to get more accurate estimates of the excess mortality attributed to the Covid-19 pandemic, as well as the difference in excess mortality between men and women in Sweden. In addition, we estimate life expectancy based on the weekly death rates. By this we aim to translate the current effect of excess mortality on total mortality, and thus provide a quantitative measure for the overall health burden.

## METHODS

### Data

Our analysis rests on weekly death counts for all-cause mortality by five-year age groups and sex from 2015 to the most recent weeks in 2020. We used monthly population counts by single years of age to calculate the respective denominator. The data is provided by Statistics Sweden. Age is measured as “highest age attained” during the respective year, and thus, death and population counts correspond to five consecutive birth cohorts. This data structure is a prerequisite to conduct mortality analysis on such detailed time units. The single-year population counts have been aggregated to match the age grouping of the death counts.

By comparing the updated death counts that are published each Monday across several weeks, we found a registration delay of around three weeks. After that time, the reported counts for the respective weeks change only marginally. Therefore, we focus our analysis on calendar weeks 10 to 16. For some deaths, the actual weeks of occurrence is reported as unknown. The amount of this deaths varies by age and sex with highest proportions of up to 10% at some young adult ages in recent years for women. These deaths have been excluded from the analysis.

### Disaggregating monthly population estimates

In order to calculate week-specific mortality measures, population counts must be disaggregated from a monthly to a weekly basis. This has been done by a weighted-average of the population at the beginning and the end of the respective month with weights that derive from the distance in days of the specific week start or end date to the respective begin or end-month population. For instance, the start-week population in the week from January 6, 2020 to January 12, 2020 was calculated by (1-6/31) times the population at December 31, 2019 plus 6/31 times the population at January 31, 2020. End-week populations have been calculated accordingly. The corresponding person-years lived during the week are the average of the start-week and end-week population times the number of weekdays divided by the total number of days in the respective year (e.g.; 7/365).

The last available population counts at the time of analysis refer to February 29, 2020. Hence, the remaining start- and end-week populations are based on extrapolations using the observed death counts only, and thus, in this number the potential effect of migration is neglected.

### Mortality measures

Our analysis focuses on the ages 50+. We started our analysis by investigating the distributions of observed age-specific death counts by week. The reference in this comparison is the range of death counts observed in the same week and the same age-group between 2015 and 2019.

In a next step, we calculated age-specific death rates on a weekly basis and estimated rate ratios between the observed death rates in 2020 and the median average death rates between 2015 and 2019. The necessary statistical inference measures have been calculated using a bootstrap procedure with Poisson distributed death counts [5].

Finally, we calculated weekly life expectancy to summarize mortality across age. This has been done for each year and week, separately. The comparison reference is also the range of life expectancies observed in the respective week between 2015 and 2019. We use life expectancy purely as a mortality summary measure because the measure itself and its magnitude in recent years is well known to many people. Thus, we do not intent to use this measure to draw any long-term conclusions. For the transformation of age-specific death rates into probabilities of dying, we borrowed information on average person-years lived by those who die within an age interval from Swedish life tables published by the Human Mortality Database [6]. The respective values correspond to the age-specific value in the life table of the year with the most similar death rate in that age group. Furthermore, the estimation of life expectancy based on small population numbers, as is the case for both weekly death counts and weekly estimates of person-years lived, can be problematic. Previous studies concluded that it is not recommended to calculate the measure for populations with less than 5,000 person-years under risk [7], However, the total number of weekly person-years under risk in our analysis is way above this threshold.

## RESULTS

Figure 1 presents the distribution of deaths over age (50 to 100+) for men and women in 2020 (points) and range of observed counts between the years 2015 to 2019 (shaded area) for calendar weeks 10 to 16 (March to mid April). Note that Sweden reached 1000 officially confirmed Covid-19 cases during week 11, 5000 confirmed cases during week 14,10000 confirmed cases during week 15 and almost 15000 confirmed cases at the end of week 16 (Source: https://coronavirus.ihu.edu/map.html).

**Fig. 1.**
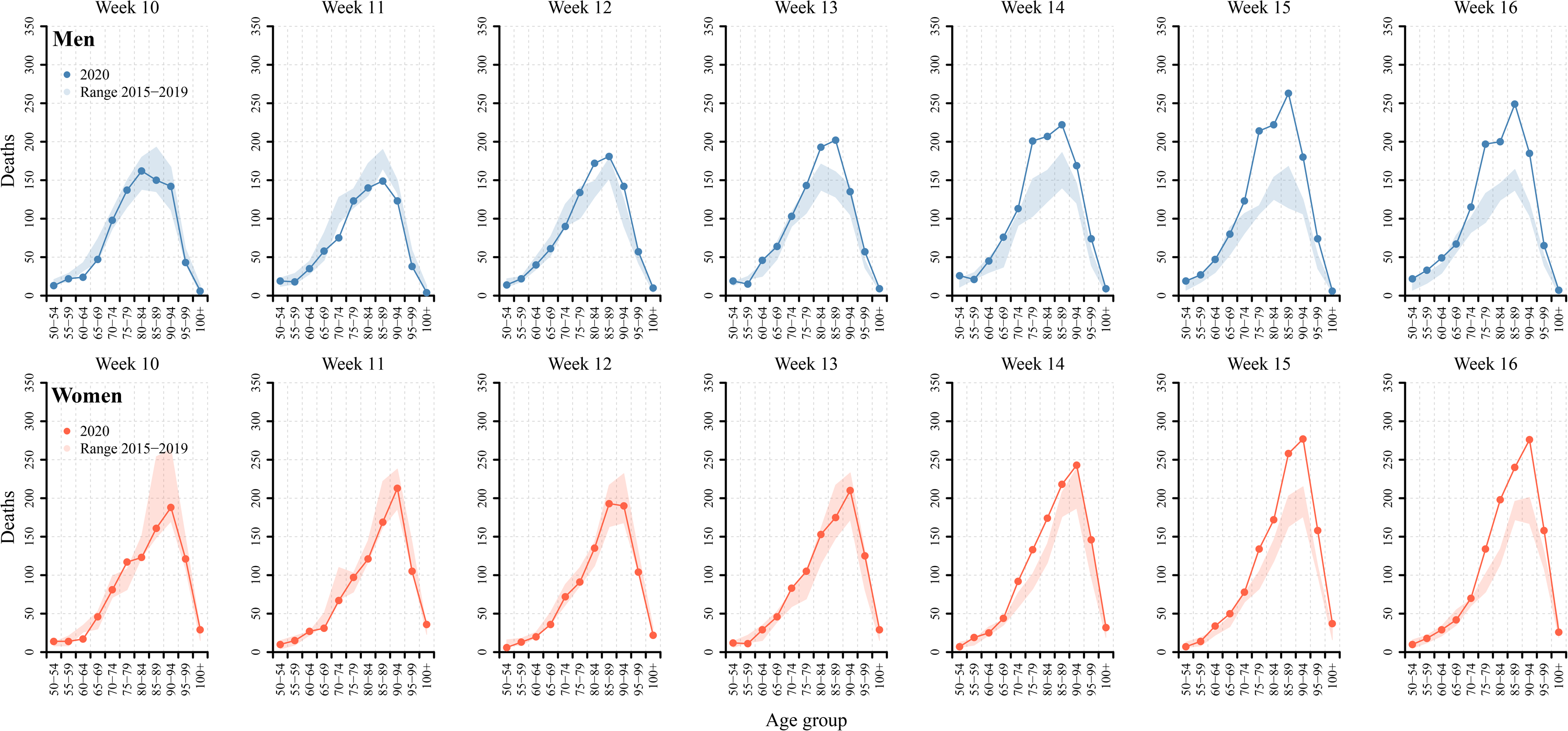
Distribution of deaths over five years age-groups from ages 50 to 100+ for weeks 10-16 in 2020, and the corresponding weeks in the previous five years, men and women, Sweden. Notes: The shaded area corresponds the range of observed death counts between 2015 and 2019. The definition of age corresponds to “highest age-attained during the respective year”, and thus, corresponds to death counts measured for birth cohorts.

The distribution of death counts over age kept approximately the same shape but shows clear levels of excess death counts with advancing pandemic. For men, this is evident from week 13 and onwards, and for women from week 14 and onwards. It is clear that the excess mortality in 2020 compared to the comparison period is larger for men than women. The figures also show that excess death counts can be found in a broader range for men compared to women.

Figure 2 (men) and Figure 3 (women) presents age- and sex-specific weekly death rates in 2020 (points) compared to the 95% confidence interval of average death rates for the corresponding weeks from 2015-2019 (shaded area). The lower panels depict rate ratios of the death rates in 2020 compared to the median average death rate of the previous years. Until week 12, mortality in 2020 was lower compared to the previous years for both men and women. However, from week 13 onward, death rates in 2020 started to exceed the previous years. This pattern becomes more and more pronounced with advancing pandemic. From the rate ratios, it becomes apparent that the highest age groups, i.e. ages 80 and above, are dis-proportionally stronger affected by the pandemic. For instance, in week 15, individuals at ages 85 to 99 are exposed to death rates that are 50% to 70% higher than the median average death rates for the years 2015 to 2019, while individuals between ages 60 to 79 are exposed to death rates that are around 25% higher. The rate ratios also show that excess mortality is generally higher among men than women.

**Fig 2.**
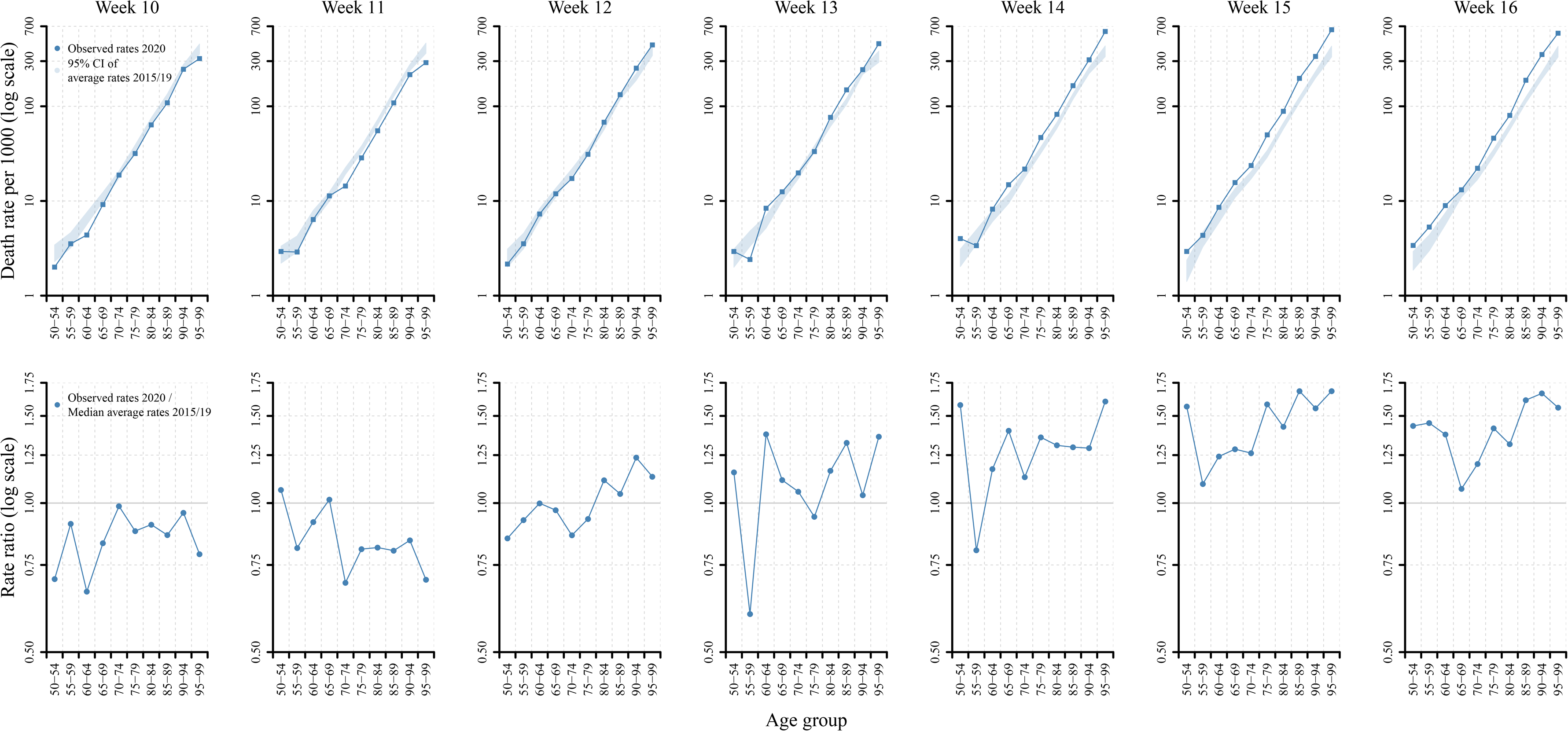
Age and sex specific weekly death rates in 2020 compared to the 95% confidence interval of average death rates for the corresponding weeks in 2015-2019, as well as the rate ratio of the death rates in 2020 compared to median average death rates for the previous years, men. Notes: The shaded are is showing the 95% confidence intervals of age-specific average death rates for the period 2015 to 2019. The interval has been calculated based on bootstrapping procedure. From the same calculations, we also derived the median age-specific average death rates that serve as the reference for the rate ratios in the lower panel.

**Fig 3.**
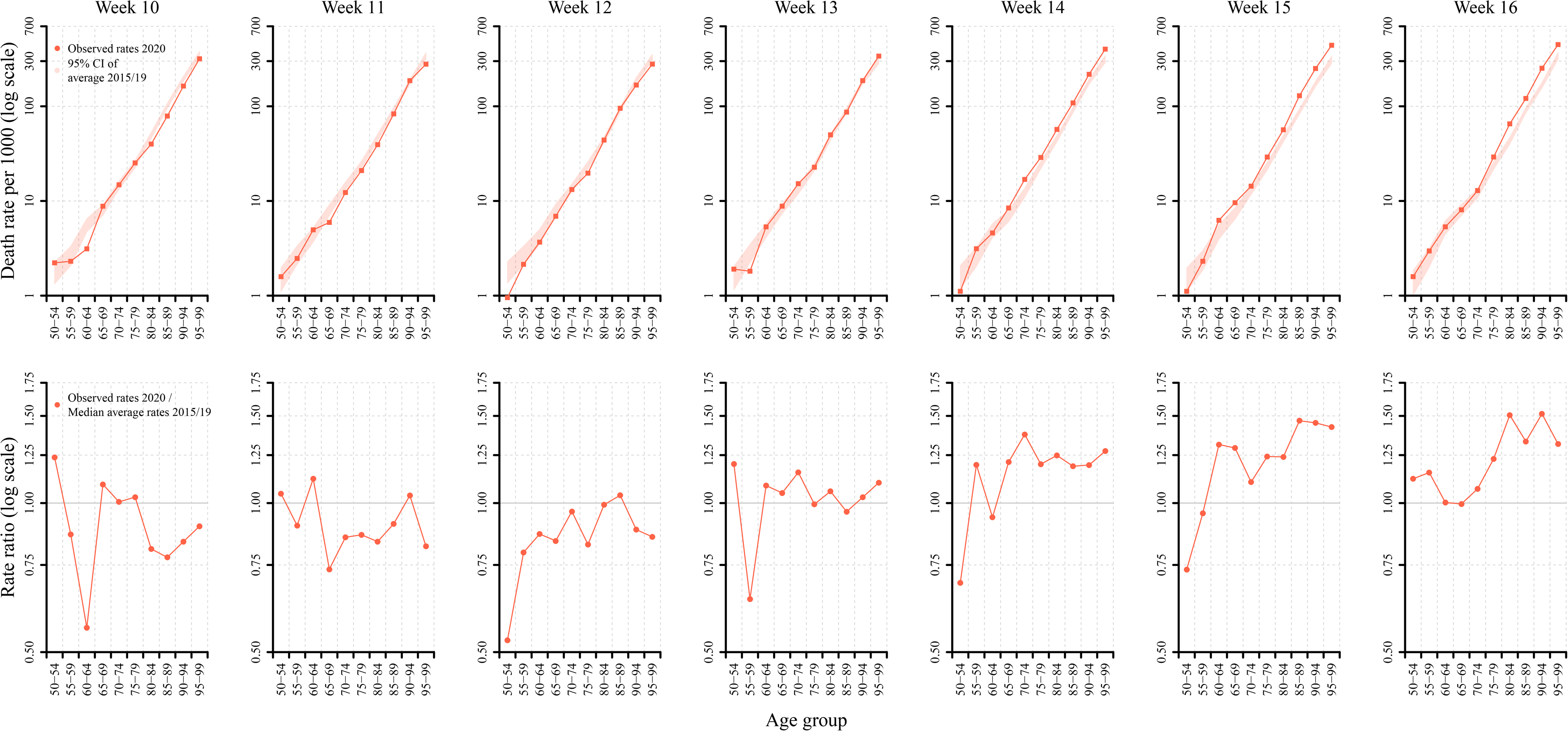
Age and sex specific weekly death rates in 2020 compared to the 95% confidence interval of average death rates for the corresponding weeks in 2015-2019, as well as the rate ratio of the death rates in 2020 compared to median average death rates for the previous years, women. Notes: The shaded area is showing the 95% confidence intervals of age-specific average death rates for the period 2015 to 2019. The interval has been calculated based on bootstrapping procedure. From the same calculations, we also derived the median age-specific average death rates that serve as the reference for the rate ratios in the lower panel.

Figure 4 shows the remaining life expectancy per week at age 50 and the 95% confidence interval for men and women in Sweden in 2020 (blue and red curve and corresponding shaded area) and the range of observed life expectancy values between 2015 and 2019 (grey area). The seasonal pattern in life expectancy is visible for the 2015-2019 period with lower life expectancy during the winter months and higher life expectancy during the summer months. This general pattern suggests that in absence of Covid-19, life expectancy would start to increase around the calendar weeks 10 to 16 due to lower mortality during the late spring and summer months. However, the excess mortality in 2020 has caused a clear drop in life expectancy. With a drop from around 33.5 years prior to the pandemic to around 30.5 in week 15, the decline amounts to 3 years for men. The corresponding numbers for women are around 36 years prior to the pandemic and 34 years in week 15. Hence, the decline for women amounts to around 2 years. However, it is important to note that this numbers are snap shots estimated for the 6 weeks in 2020. They are therefore only showing current mortality levels and not the final impact of Covid-19 on the level of life expectancy. However, these patterns and the observation that the levels of life expectancy in week 15 correspond to the annual levels observed in 2006 for Swedish men and women [6] gives a perception about the magnitude of excess mortality from the Covid-19 pandemic on total mortality in Sweden.

**Fig 4.**
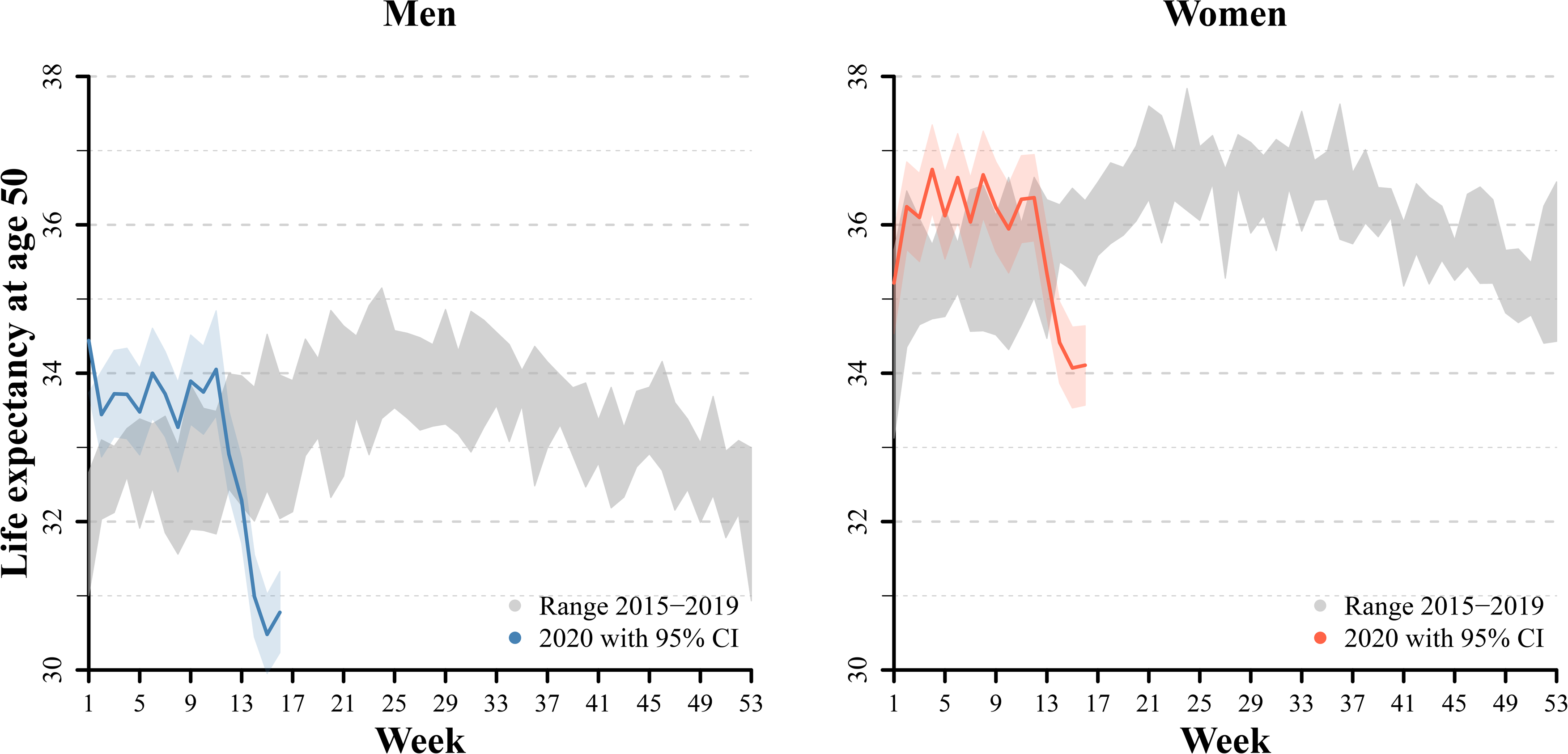
Median estimate of remaining life expectancy at age 50 per calendar week for men and women in 2020 with 95% confidence interval compared to the range of observed weekly life expectancies between 2015 to 2019. Notes: The shaded area is showing the 95% confidence intervals of weekly life expectancy in 2020, while the solid line depicts the corresponding median estimate. The interval has been calculated based on a bootstrapping procedure that we applied to the age-specific death counts.

## DISCUSSION

The consequences that the Covid-19 pandemic will have on total mortality and life expectancy in Sweden and other countries cannot be known at this point in time. However, by following weekly all-cause death rates and compare them to previous years, we can get a perception of the health burden. Our analysis for Sweden showed that from week 13 and onwards (the last week of march) mortality started to increase. Two weeks earlier (week 11), Sweden surpassed the number of 1000 confirmed Covid-19 cases. From week 14 and onwards, the death rates at all ages above 60 are higher than those in previous years. The general pattern of excess mortality compared to previous years is clear and consistent. The pattern shows also that persons above age 80 are dis-proportionally stronger affected and also that men suffer higher levels of excess mortality than women. To get an idea of the overall magnitude of excess deaths on total mortality, we estimated life expectancy per week and showed that the excess mortality in 2020 corresponds to a decline in life expectancy of 3 years for men and 2 years for women during week 13 to week 16. This is in line with a recently published working paper by Goldstein and Lee [8], who estimated, among other things, the reduction in remaining life expectancy to 3 years for the US, and a recent preprint paper from Italy estimating the life expectancy impact for northern Italy at a similar range [9].

The mortality changes occurred over a period of 13 weeks after Sweden’s first confirmed case of Covid-19 (Jan 31^st^) and 2-3 weeks after community transmission was confirmed (March 9^th^) in the Stockholm region. Sweden has so far taken a somewhat different approach than most countries for the containment of the virus. The strategy is built on recommendations and that individuals take their own responsibility for social distancing, and without a strict lockdown. The strategy has, at least indirectly, suggested to aim for herd immunity at a controlled pace. However, together with Italy, Spain, France, the UK and Belgium, Sweden is among the countries with highest reported excess mortality (Source: https://euromomo.eu/). If the pandemic will eventually also lead to declining life expectancy for the entire calendar year 2020 will depend on the progression of the pandemic, the extent that some of the deaths would have occurred regardless of Covid-19, only somewhat earlier than they otherwise would have, the consequences for other health conditions, as well as the health care sector at large. With other health conditions, we mean for example potentially raised mortality due to changes in the utilization of emergency care, prolonged waiting times, or raised psychiatric illness and suicide due to for instance social isolation. It has been hypothesized that the effect of the pandemic will not have a huge impact on life expectancy since many deaths in old age and risk groups would have occurred even in the absence of Covid-19, only somewhat later. However, a recent study suggests the opposite and conclude that virus can be harmful for individuals at all levels of frailty and that it has cut lifespans be at least a decade on average [10]. The death toll we found is in line with this finding and suggest at least the number of deaths in near future must reach unrealistically low levels to off-set the tragic losses observed until recently.

Our results also confirm the sex difference in mortality that are based on death counts and reported since the outbreak of Covid-19. Although subject to fluctuations, the rate ratios of the age-specific death rates show approximately similar general age patterns for men and women. However, the magnitude of excess mortality is higher for men at all ages. We cannot draw any conclusions about whether the mortality difference truly stem from different chances of surviving Covid-19, or a difference in the risk of getting affected, neither can we explain the mechanisms to the observed sex differences. Even if the number of confirmed cases is similar for men and women in Sweden, and in other countries, it is still confirmed cases and not all cases. However, based on the confirmed cases it clearly seems as if men have higher fatality. This together with our finding that excess mortality follows the same age-pattern for men and women but with higher levels for men at all ages may suggest biological explanations. This finding relates to a preprint paper that reports the same susceptibility to Covid-19 for men and women but that men seem to be more prone to have higher severity and fatality independent of age [5].

## CONCLUSION

From the first week of April and onwards, the death rates at all ages above 60 are higher than those in previous years in Sweden. The pattern shows that persons above age 80 are dis-proportionally more affected, and that men suffer higher levels of excess mortality than women. Currently this excess mortality corresponds to a decline in remaining life expectancy of 3 and 2 years for men and women respectively. What consequences the pandemic will eventually have on life expectancy will be determined by a complex interplay of several factors. Until now, the Covid-19 pandemic however has already caused a health burden that reached historic extents on the total mortality of the Swedish population.

## Data Availability

The data underlying this study can be receievd from Statistics Sweden.

## ACKNOWLEDGEMENTS

We thank Statistic Sweden and Tomas Johansson for providing us with data, and Professor Anders Ahlbom, Karolinska Institutet who provided useful comments on earlier versions of this manuscript.

## Contributors

KM and ME conceived and designed the study. KM obtained access to data. ME conducted the data analysis. KM drafted the initial version of the manuscript. KM and ME together interpreted the data and critically revised the manuscript. Both authors had full access to data and figures in the study and can take responsibility for the integrity of the data and the accuracy of the data analysis. KM is the guarantor. The corresponding author attests that all listed authors meet authorship criteria and that no others meeting the criteria have been omitted.

## Ethical approval

Not applicable.

## REFERENCES

1. Statista. Coronavirus (COVID-19) death rate in countries with confirmed deaths and over 1000 reported cases as of April 30 2020, by country. https://www.statista.com/statistics/1105264/coronavirus-covid-19-cases-most-affected-countries-worldwide/

2. Dudel, C., Riffe, T., Acosta, E., van Raalte, A. A., Strozza, C., & Myrskylä, M., Monitoring trends and differences in COVID-19 case fatality rates using decomposition methods: Contributions of age structure and age-specific fatality. 2020. SocArXiv Preprint

3. Leon, D.A., et al., COVID-19: a need for real-time monitoring of weekly excess deaths. Lancet, 2020. 395(10234): p. e81.

4. Hannah Peckham, N.d.G., Charles Raine et al., Sex-bias in COVID-19: a meta-analysis and review of sex differences in disease and immunity, 20 April 2020, PREPRINT (Version 2) available at Research Square [+https://doi.ora/10.21203/rs.3.rs-23651.2020.

5. Brillinger, D.R., A Biometrics Invited Paper with Discussion: The Natural Variability of Vital Rates and Associated Statistics. Biometrics, 1986. 42(4): p. 693–734.

6. Human Mortality Database. University of California, B.U., and Max Planck Institute for Demographic Research (Germany). Available at www.mortalitv.org.

7. Eayres, D. and E.S. Williams, Evaluation of methodologies for small area life expectancy estimation. Journal of epidemiology and community health, 2004. 58(3): p. 243–249.

8. Goldstein, J.R. and Lee, R.D., Demographic Perspectives on Mortality of Covid-19 and Other Epidemics. NBER WORKING PAPER SERIES. Working Paper 27043. April 2020.

9. Ghislandi, S., et al., News from the front: Excess mortality and life expectancy in two major epicentres of the COVID-19 pandemic in Italy. Preprint medRxiv, 2020: p. 2020.04.29.20084335.

10. Hanlon, P., et al., COVID-19 ? exploring the implications of long-term condition type and extent of multimorbidity on years of life lost: a modelling study [version 1; peer review: awaiting peer review], Wellcome Open Research, 2020. 5(75).

